# NeuroMark: a fully automated ICA method to identify effective fMRI markers of brain disorders

**DOI:** 10.1101/19008631

**Authors:** Yuhui Du, Zening Fu, Jing Sui, Shuang Gao, Ying Xing, Dongdong Lin, Mustafa Salman, Md Abdur Rahaman, Anees Abrol, Jiayu Chen, L Elliot Hong, Peter Kochunov, Elizabeth A. Osuch, Vince D. Calhoun, for the Alzheimer’s Disease Neuroimaging Initiative

## Abstract

Increasing sharing initiatives on neuroimaging data have provided unprecedented opportunities to study brain disorders. Standardized approaches for capturing reproducible and comparable biomarkers are greatly needed. Here, we propose a method, *NeuroMark*, which leverages a priori-driven independent component analysis to effectively extract functional brain network biomarkers from functional magnetic resonance imaging (fMRI) data. *NeuroMark* automatically estimates features adaptable to each individual and comparable across subjects by taking advantage of the replicated brain network templates extracted from 1828 healthy controls as guidance to initialize the individual-level networks. Four studies including 2454 subjects were conducted spanning six brain disorders (schizophrenia, autism spectrum disorder, depression, bipolar disorder, mild cognitive impairment and Alzheimer’s disease) to evaluate the proposed method from different perspectives (replication, cross-study comparison, subtle difference identification, and multi-disorder classification). Results demonstrate the great potential of *NeuroMark* in its feasibility to link different datasets/studies/disorders and enhance sensitivity in identifying biomarkers for patients with challenging mental illnesses.

**Significance Statement:** Increasing evidence highlights that features extracted from resting fMRI data can be leveraged as potential biomarkers of brain disorders. However, it has been difficult to replicate results using different datasets, translate findings across studies, and differentiate brain disorders sharing similar clinical symptoms. It is important to systematically characterize the degree to which unique and similar impaired patterns are reflective of brain disorders. We propose a fully automated method (called *NeuroMark*) that leverages priori-driven independent component analysis (ICA) using replicated brain network templates to estimate individual-subject network features. Evaluated by four studies involving six different brain disorders, we show that *NeuroMark* can effectively link the comparison of biomarkers across different studies/datasets/disorders and enable classification between complex brain disorders, while also providing information about relevant aspects of whole brain functional connectivity.

## Introduction

In the neuroscience field, increasing data-sharing initiatives have accelerated the use of neuroimaging to study brain disorders in the clinic (Poldrack and Gorgolewski, 2014; Poline et al., 2012; Woo et al., 2017). Access to multi-site datasets affords unprecedented opportunities to perform large-scale analysis across disorders. However, systematic methods to estimate neuroimaging measures in the context of enhancing neuroscientific validity are still very limited. There is a need for techniques that can accelerate the identification of interpretable brain markers in preexisting data and evaluation of biomarkers in terms of their generalizability, reproducibility and relationship to other data.

Many approaches have been utilized to capture neuroimaging features informative of brain functional connectivity from functional magnetic resonance imaging (fMRI) data, including region of interest (ROI) or seed derived connectivity analysis (Dosenbach et al., 2010; Tzourio-Mazoyer et al., 2002), self-activation detection method (such as amplitude of low frequency fluctuation and regional homogeneity (Zang et al., 2004)), decomposition-based independent component analysis (ICA) (Calhoun and Adali, 2012; Calhoun et al., 2001; McKeown et al., 2003), as well as clustering techniques to group brain voxels (Du et al., 2012). ROI analysis and ICA are two of the most commonly used approaches for studying functional connectivity. While ROI-based methods typically require fixed brain regions according to prior experience or knowledge, ICA, a data-driven method, is capable of capturing functional networks while retaining more single-subject variability (Yu et al., 2017). ICA leverages the hidden spatio-temporal information to extract maximally spatially independent components (ICs), each of which includes brain voxels sharing co-varying patterns. Other advantages of ICA are that it can perform intrinsic connectivity network (ICN) extraction and noise component removal simultaneously (Du et al., 2016a), while also enabling separation of overlapping but distinct functional activity (Xu et al., 2015). However, blind ICA is challenging for multi-subject data analyses, since components extracted from different subjects may not have spatial correspondence. To overcome the ICA correspondence limitation, we and others have developed group ICA methods (Beckmann et al., 2009; Calhoun et al., 2001; Calhoun and de Lacy, 2017; Du and Fan, 2013). The majority of these approaches involve an ICA performed on the group data to estimate the group-level components, and then utilize a back-reconstruction strategy to extract individual-level functional networks and associated time-courses.

Group ICA still has limitations because comparing results from different group ICAs is not straightforward, which affects its ability to replicate findings from different studies. For example, a study (Allen et al., 2014) identified 50 ICNs arranged into seven functional domains, and another study (Marusak et al., 2017) characterized 52 ICNs sorted to three domains, despite using the same model order. Such differences in the identified ICNs and their arrangements hinder the direct comparisons across the results. The non-correspondence between group ICA runs also leads to a problem in studies focusing on classification where group ICA is usually performed on all subjects’ fMRI data to make the resulting features work for the trained classifier (Demirci et al., 2008; Rashid et al., 2016). This operation can be biased, as the feature extraction should be independent from the testing data.

Accordingly, it is essential to adopt an ICA approach to estimate brain network measures in an unbiased way that fully persist individual property while also being able to be compared across subjects from various studies.

From a clinical perspective, an analysis method that can link different brain disorders is greatly needed so as to evaluate sensitivity and specificity as well as similarity and overlap between disorders using neuroimaging data. Although some brain disorders can be assigned to distinct categories, many share clinically-overlapping symptoms. For example, both schizoaffective and bipolar disorders experience hallucinations and delusions that are typical features of schizophrenia (SZ) (Cosgrove and Suppes, 2013), which can make their clinical differentiation difficult. As such, it is beneficial to investigate shared and unique brain impairments among these symptom-related disorders. Furthermore, it may be promising to develop new biologically-based types across the psychotic illnesses (Colibazzi, 2014) by combining the use of image-derived features. Effective method will accelerate the refinement of current disorder categories using consistent neural measures. Taking SZ and autism spectrum disorder (ASD) into account, although they are conceptualized as distinct illnesses, they have also been revisited in recent years due to their shared phenotypic and genotypic expression (Hommer and Swedo, 2015). Unfortunately, there is a paucity of studies that perform a direct comparison of symptom-related disorders and the validation of brain changes using large-sample datasets, probably due to the limited ability of analytic methods to characterize individual variability (Zuo et al., 2014) and reliability (Noble et al., 2019). Since more neuroimaging data are now available than ever before, we have an opportunity to probe this aim. A method that can optimize the data-specific variability while retaining the comparability across different datasets, studies and disorders is urgently needed.

To address the very important problem, we design *NeuroMark*, a method that can leverage group information guided ICA (GIG-ICA) (Du and Fan, 2013) or spatially constrained ICA (Lin et al., 2010) to fully automate the estimation and labeling of individual-subject network features, by incorporating an additional input of spatial network priors derived from independent large samples. We conducted four studies involving six brain disorders and more than 2400 subject samples to validate the reliable performance of *NeuroMark* in different aspects, including the replication of biomarkers on independent datasets, the cross-study comparison for linking related disorders, the extraction of subtle group difference across progressively developing disorders, and the classification on challengeable brain disorders. Our results highlight that *NeuroMark* effectively identified replicated biomarkers of schizophrenia across different datasets; revealed interesting neural clues on the overlap between autism and schizophrenia; demonstrated dimensional brain functional impairments present to varying degrees in mild cognitive impairments and Alzheimer’s disease; and achieved high performance in classifying bipolar disorder and depression. While we focus on resting fMRI initially, *NeuroMark* can be expanded to incorporate multimodal imaging data as well. The code and templates are available online (www.yuhuidu.com and http://trendscenter.org/software).

## Results

The flowchart of *NeuroMark* is displayed in Fig. 1. First, replicated ICN templates are constructed from different groups of large-sample healthy controls (HCs). Next, using the ICN templates as the spatial network priors, GIG-ICA (Du et al., 2016a; Du and Fan, 2013) is applied to automatically estimate subject-specific functional networks and associated time-courses (TCs), due to the high reliability (Du et al., 2017) and accurate individual property (Salman et al., 2019) of brain functional networks estimated by the method. Finally, different network features, such as functional network connectivity (FNC) reflecting the interaction between networks, are computed and then evaluated.

**Fig. 1.**
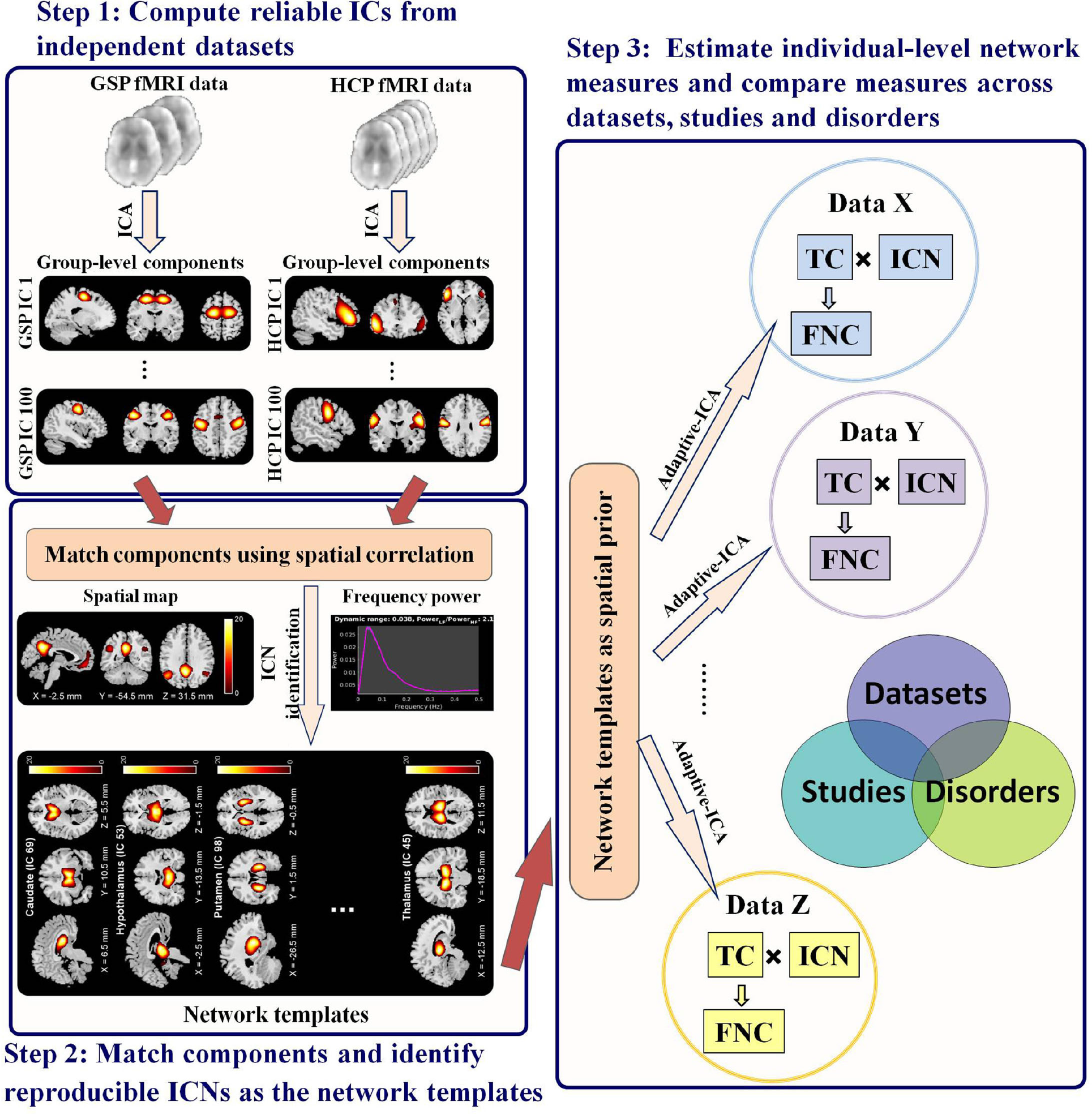
Schematic flowchart of the proposed framework. Step 1: Calculate group-level independent components (ICs) from two independent datasets with large sample of healthy controls including the human connectome project (HCP) and genomics superstruct project (GSP) datasets. Step 2: Match ICs using spatial correlations between their spatial maps and then identify highly replicated intrinsic connectivity networks (ICNs) as the network templates. Step 3: Automatically calculate the individual-level ICNs and their related time courses (TCs) by taking the network templates as prior information in Adaptive-ICA. Other extended features such as functional network connectivity (FNC) can be obtained and then compared across datasets, studies, and disorders.

We obtained 53 replicated network templates (Fig. 2) that are common between 823 HCs in the human connectome project (HCP) and 1005 HCs in the genomics superstruct project (GSP) datasets. They were arranged into seven functional domains according to their functional and anatomical roles (Allen et al., 2014), including the sub-cortical (SC: 5 ICNs), auditory (AU: 2 ICNs), sensorimotor (SM: 9 ICNs), visual (VI: 9 ICNs), cognitive control (CC: 17 ICNs), default mode (DM: 7 ICNs) and cerebellar (CB: 4 ICNs) domains. The detailed component labels and peak coordinates are provided in Table 1. We evaluated the method using four studies, showing that *NeuroMark* provides an effective way to identify and compare brain biomarkers.

**Table 1.** Information of the extracted network templates. For each template, its functional domain, primary brain region and peak coordinate are included. Here, each network template is represented by one independent component (IC). IC ID is shown along with the brain region name.

| Primary regions in ICNs (IC ID) | X | Y | Z | Primary regions in ICNs (IC ID) | X | Y | Z |
| --- | --- | --- | --- | --- | --- | --- | --- |
| **Sub-cortical domain (SC)** |  |  |  | **Cognitive-control domain (CC)** |  |  |  |
| Caudate (IC 69) | 6.5 | 10.5 | 5.5 | Inferior parietal lobule ([IPL], IC 68) | 45.5 | -61.5 | 43.5 |
| Subthalamus/hypothalamus (IC 53) | -2.5 | -13.5 | -1.5 | Insula (IC 33) | -30.5 | 22.5 | -3.5 |
| Putamen (IC 98) | -26.5 | 1.5 | -0.5 | Superior medial frontal gyrus ([SMFG], IC 43) | -0.5 | 50.5 | 29.5 |
| Caudate (IC 99) | 21.5 | 10.5 | -3.5 | Inferior frontal gyrus ([IFG], IC 70) | -48.5 | 34.5 | -0.5 |
| Thalamus (IC 45) | -12.5 | -18.5 | 11.5 | Right inferior frontal gyrus ([R IFG], IC 61) | 53.5 | 22.5 | 13.5 |
| **Auditory domain (AU)** |  |  |  | Middle frontal gyrus ([MiFG], IC 55) | -41.5 | 19.5 | 26.5 |
| Superior temporal gyrus ([STG], IC 21) | 62.5 | -22.5 | 7.5 | Inferior parietal lobule ([IPL], IC 63) | -53.5 | -49.5 | 43.5 |
| Middle temporal gyrus ([MTG], IC 56) | -42.5 | -6.5 | 10.5 | Left inferior parietal lobue ([R IPL], IC 79) | 44.5 | -34.5 | 46.5 |
| **Sensorimotor domain (SM)** |  |  |  | Supplementary motor area ([SMA], IC 84) | -6.5 | 13.5 | 64.5 |
| Postcentral gyrus ([PoCG], IC 3) | 56.5 | -4.5 | 28.5 | Superior frontal gyrus ([SFG], IC 96) | -24.5 | 26.5 | 49.5 |
| Left postcentral gyrus ([L PoCG], IC 9) | -38.5 | -22.5 | 56.5 | Middle frontal gyrus ([MiFG], IC 88) | 30.5 | 41.5 | 28.5 |
| Paracentral lobule ([ParaCL], IC 2) | 0.5 | -22.5 | 65.5 | Hippocampus ([HiPP], IC 48) | 23.5 | -9.5 | -16.5 |
| Right postcentral gyrus ([R PoCG], IC 11) | 38.5 | -19.5 | 55.5 | Left inferior parietal lobue ([L IPL], IC 81) | 45.5 | -61.5 | 43.5 |
| Superior parietal lobule ([SPL], IC 27) | -18.5 | -43.5 | 65.5 | Middle cingulate cortex ([MCC], IC 37) | -15.5 | 20.5 | 37.5 |
| Paracentral lobule ([ParaCL], IC 54) | -18.5 | -9.5 | 56.5 | Inferior frontal gyrus ([IFG], IC 67) | 39.5 | 44.5 | -0.5 |
| Precentral gyrus ([PreCG], IC 66) | -42.5 | -7.5 | 46.5 | Middle frontal gyrus ([MiFG], IC 38) | -26.5 | 47.5 | 5.5 |
| Superior parietal lobule ([SPL], IC 80) | 20.5 | -63.5 | 58.5 | Hippocampus ([HiPP], IC 83) | -24.5 | -36.5 | 1.5 |
| Postcentral gyrus ([PoCG], IC 72) | -47.5 | -27.5 | 43.5 | **Default-mode domain (DM)** |  |  |  |
| **Visual domain (VI)** |  |  |  | Precuneus (IC 32) | -8.5 | -66.5 | 35.5 |
| Calcarine gyrus ([CalcarineG], IC 16) | -12.5 | -66.5 | 8.5 | Precuneus (IC 40) | -12.5 | -54.5 | 14.5 |
| Middle occipital gyrus ([MOG], IC 5) | -23.5 | -93.5 | -0.5 | Anterior cingulate cortex ([ACC], IC 23) | -2.5 | 35.5 | 2.5 |
| Middle temporal gyrus ([MTG], IC 62) | 48.5 | -60.5 | 10.5 | Posterior cingulate cortex ([PCC], IC 71) | -5.5 | -28.5 | 26.5 |
| Cuneus (IC 15) | 15.5 | -91.5 | 22.5 | Anterior cingulate cortex ([ACC], IC 17) | -9.5 | 46.5 | -10.5 |
| Right middle occipital gyrus ([R MOG], IC 12) | 38.5 | -73.5 | 6.5 | Precuneus (IC 51) | -0.5 | -48.5 | 49.5 |
| Fusiform gyrus (IC 93) | 29.5 | -42.5 | -12.5 | Posterior cingulate cortex ([PCC], IC 94) | -2.5 | 54.5 | 31.5 |
| Inferior occipital gyrus ([IOG], IC 20) | -36.5 | -76.5 | -4.5 | **Cerebellar domain (CB)** |  |  |  |
| Lingual gyrus ([LingualG], IC 8) | -8.5 | -81.5 | -4.5 | Cerebellum ([CB], IC 13) | -30.5 | -54.5 | -42.5 |
| Middle temporal gyrus ([MTG], IC 77) | -44.5 | -57.5 | -7.5 | Cerebellum ([CB], IC 18) | -32.5 | -79.5 | -37.5 |
|  |  |  |  | Cerebellum ([CB], IC 4) | 20.5 | -48.5 | -40.5 |
|  |  |  |  | Cerebellum ([CB], IC 7) | 30.5 | -63.5 | -40.5 |

**Fig. 2.**
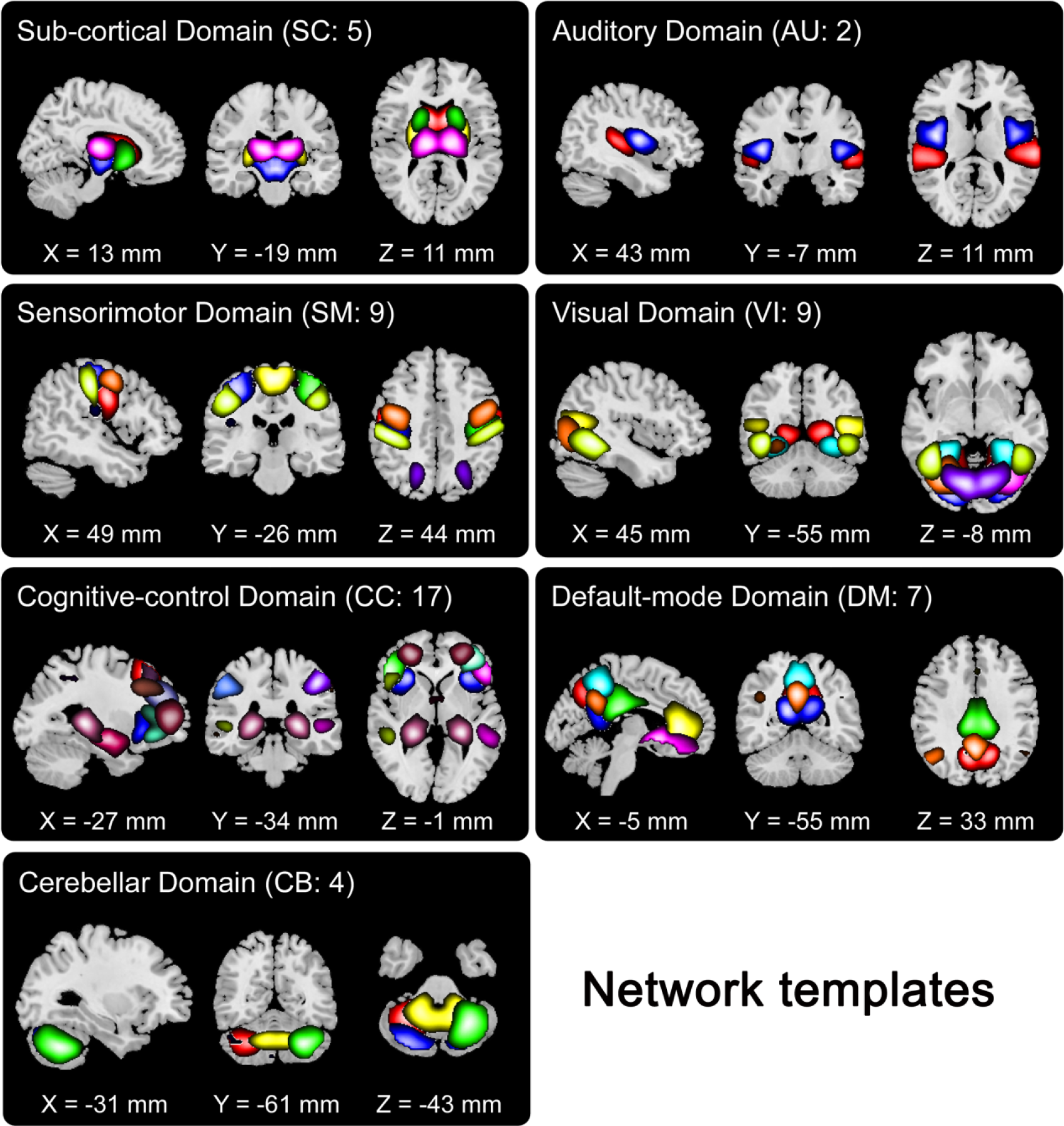
Visualization of the identified network templates, which were divided into seven functional domains based on their anatomical and functional properties. In each subfigure, one color in the composite maps corresponds to an ICN.

### Study 1: SZ patients show significant and replicable static FNC alterations using different datasets

In the study, we assess the ability of our method to identify reproducible biomarkers of schizophrenia. We used the Function Biomedical Informatics Research Network (FBIRN) dataset including 137 SZ patients and 144 HCs and another dataset collected at the University of Maryland, Maryland Psychiatric Research Center (MPRC) including 150 SZ patients and 238 HCs. For each subject, a static FNC (sFNC) matrix was obtained by computing Pearson correlation between the time series of any two ICNs. The sFNC pattern showed consistency between the FBIRN and MPRC datasets (see Fig. 3(A) and (D)), indicating the comparability of network features computed with the guidance of templates. For FBIRN, we evaluated the differences between HC and SZ by performing two-sample t-test on each connectivity related values in the sFNC matrices. Fig. 3(B) and (C) display the original T-value map for all connections and the significant group differences after the multiple comparisons correction, respectively. A similar difference pattern was found using the MPRC data, as shown in Fig. 3(E) and (F), providing evidence that the SZ-related functional impairments identified using *NeuroMark* are reproducible.

**Fig. 3.**
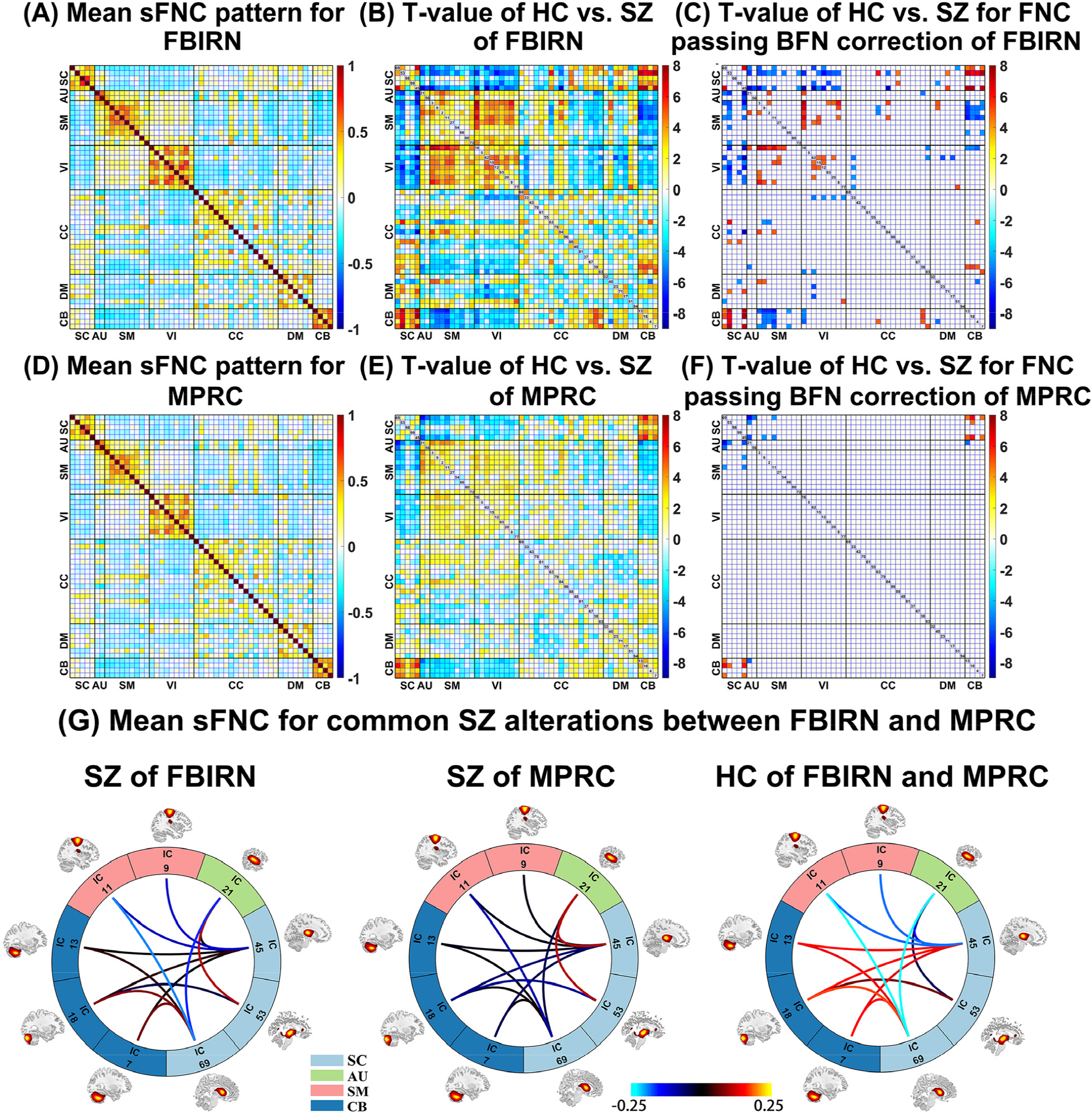
Result of study 1, which shows that there are reproducible sFNC alterations of SZ between FBIRN and MPRC data. (A) and (D): Mean sFNC pattern across all subjects for FBIRN and MPRC, respectively. (B) and (E): The T-values of all sFNCs from two-sample t-tests for FBIRN and MPRC, respectively. (C) and (F): The T-values for the remaining sFNCs after the multiple comparisons correction (p <0.05 with Bonferroni correction) for FBIRN and MPRC, respectively. “BFN” denotes Bonferroni correction. (G): The mean sFNC strength across subjects for the HC and SZ groups in the common impairments between FBIRN and MPRC data. For each commonly changed sFNC, the averaged values in SZ patients of FBIRN, SZ patients of MPRC, and HCs of the two datasets are shown, respectively.

Regarding both datasets, the primary differences in SZ compared to HC were consistently located in the connectivity between the SC and CB domains, between the SC and AU domains, as well as between the SC and SM domains. Furthermore, the common significantly changed functional connectivity was evaluated based on Fig. 3(C) and (F). Compared to HC, SZ showed decreased connectivity strength between thalamus and cerebellum, caudate and cerebellum, subthalamus and cerebellum, but increased strengths between thalamus and postcentral gyrus, thalamus and superior temporal gyrus, caudate and postcentral gyrus, caudate and superior temporal gyrus, as well as subthalamus and superior temporal gyrus. Our results (Fig. 3(G)) also indicated that the mean connectivity strength value of SZ patients was close between FBRIN and MPRC, although FBIRN data slightly showed enhanced connection strengths than MPRC data. Taken together, our results suggest that the *NeuroMark* method is promising in identifying reproducible network markers of brain disorders.

### Study 2: ASD and SZ show similar cross-disorder sFNC changes

In this study, we highlight the potential of *NeuroMark* in linking results across separate analyses so as to compare different brain disorders. Similar to the procedure in study 1, we evaluated sFNC changes in ASD patients relative to healthy population using fMRI data of 398 ASD patients and 471 HCs from Autism Brain Imaging Data Exchange I (ABIDEI). It is clear that for ABIDEI data, the mean sFNC matrix computed by averaging the sFNC matrices across different subjects (Fig. 4(A)) showed a similar connectivity pattern as that from study 1, again demonstrating that the network features computed using *NeuroMark* are corresponding and comparable. Statistical analysis showed significant group differences between HC and ASD primarily involving the SC, CB, AU and SM domains, as shown in Fig. 4(B) and (C).

**Fig. 4.**
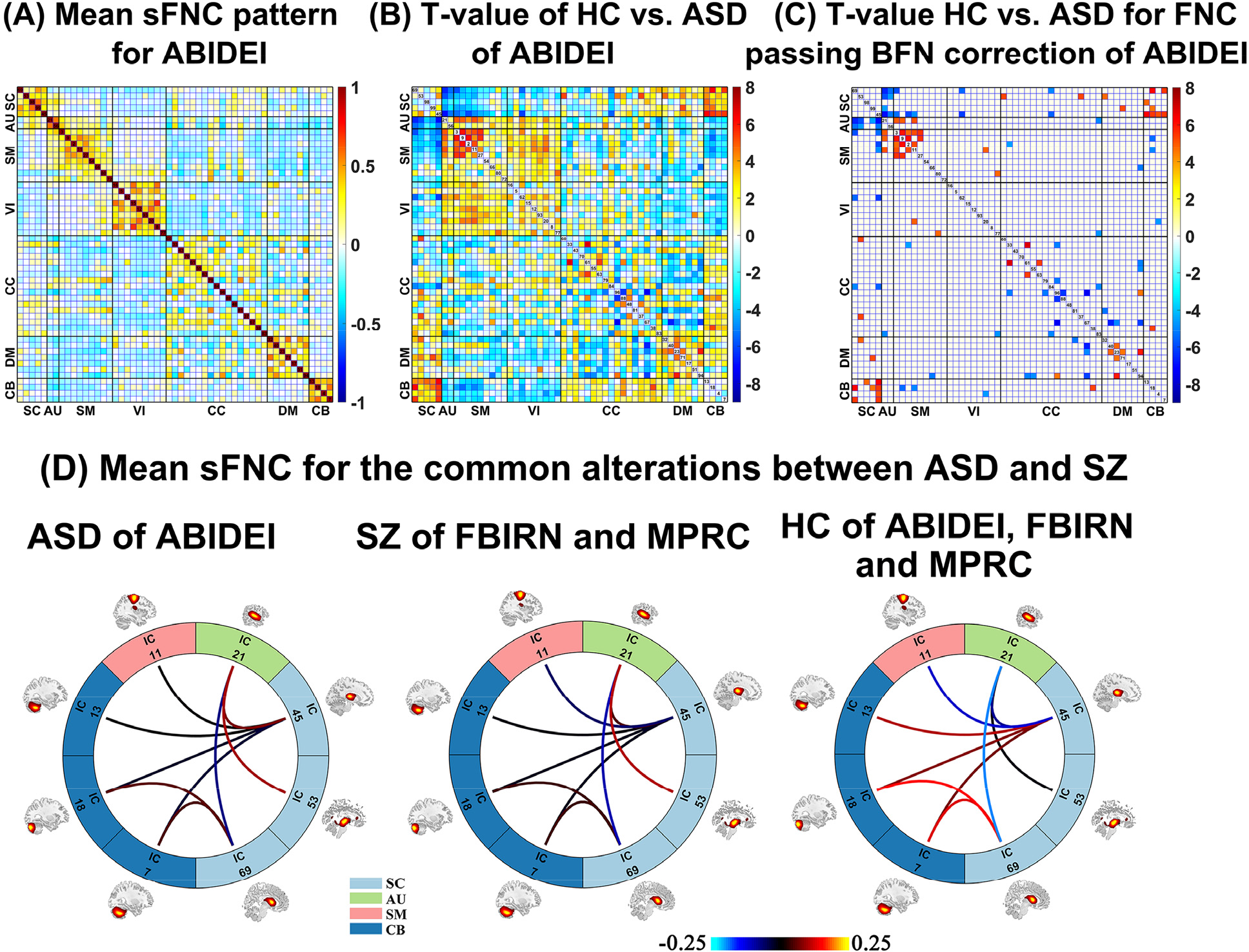
Results of study 2, which supports that SZ and ASD show common alterations in sFNC. (A): Mean sFNC pattern across all subjects for ABIDEI. (B) and (C): The T-values of all sFNCs and that of the remaining sFNCs after the multiple comparisons correction (p <0.05 with Bonferroni correction) from two-sample t-tests of HC vs. ASD for ABIDEI. “BFN” denotes Bonferroni correction. (D) Mean sFNC strength of each group (ASD, SZ and HC) in the overlapping impairments between SZ and ASD. For each commonly impaired sFNC, the averaged values in ASD patients of ABIDEI, SZ patients of FBIRN and MPRC, and HCs of the three datasets are shown, respectively.

While our findings in terms of ASD alterations could stand on its own as a result, we were interested in learning additional information by linking study 1 and the above-mentioned analysis. By comparing the common HC vs. SZ connection differences in both FBIRN and MPRC with the HC vs. ASD connection differences from ABIDEI, we identified nine overlapping sFNC alterations between SZ and ASD relative to HC. Among the nine overlaps between SZ and ASD impairments, four connections showed decrease between cerebellum and thalamus (or caudate); four connections showed increase between superior temporal gyrus and subcortical regions; and the remaining one showed increase between postcentral gyrus and thalamus. In summary, as illustrated in Fig. 4(D), the sub-cortical, sensorimotor, auditory and cerebellar regions related sFNC strengths were commonly affected for both SZ and ASD patients. Moreover, sub-cortical regions showed the largest number of common impairments across the two disorders.

### Study 3: Mild cognitive impairment (MCI) demonstrates intermediate dynamic FNC changes between HC and Alzheimer’s disease (AD)

In this study, we aim to show that *NeuroMark* can effectively capture subtle differences in dynamic functional network connectivity (dFNC) features among progressively developing brain disorders. The analysis focused on Alzheimer’s disease (AD) and mild cognitive impairment (MCI) using the Alzheimer’s Disease Neuroimaging Initiative (ADNI) dataset.

Based on fMRI data of 104 AD patients, 470 MCI patients, and 264 HCs, the dFNC patterns were first computed by a sliding time window method (Hutchison et al., 2013) using the time-series of individual-subject networks extracted by *NeuroMark*. Next, the reliable connectivity states were extracted from dFNC through a clustering technique (Allen et al., 2014). In the group-discriminating states (Fig. 5), specifically the state 2 that accounts for >50% of all windows resembled the sFNC patterns; the state 1 revealed negative connectivity strengths between SM and VI; and the state 3 in contrast showed a strong positive connection between SM and VI, suggesting that these states captured unique recurring patterns of connectivity. We found that compared to HCs, AD patients had a significantly different fraction rate of occurrences in each of the dFNC states. In general, AD patients spent less time in strongly-connected states (i.e. state 1 and state 3, which showed strong correlated and anticorrelated connectivity strengths) but more time in weakly-connected states (i.e. state 2 and state 5). Although there was no significant group difference in the state occurrences between MCI and HC/AD, MCI showed a similar but weaker changing trend with AD. Looking into this further, when separating the MCI group into early MCI (EMCI) and late MCI (LMCI), the gradually changing patterns from HC to EMCI to LMCI to AD were clearly observed (Fig. 5).

**Fig. 5.**
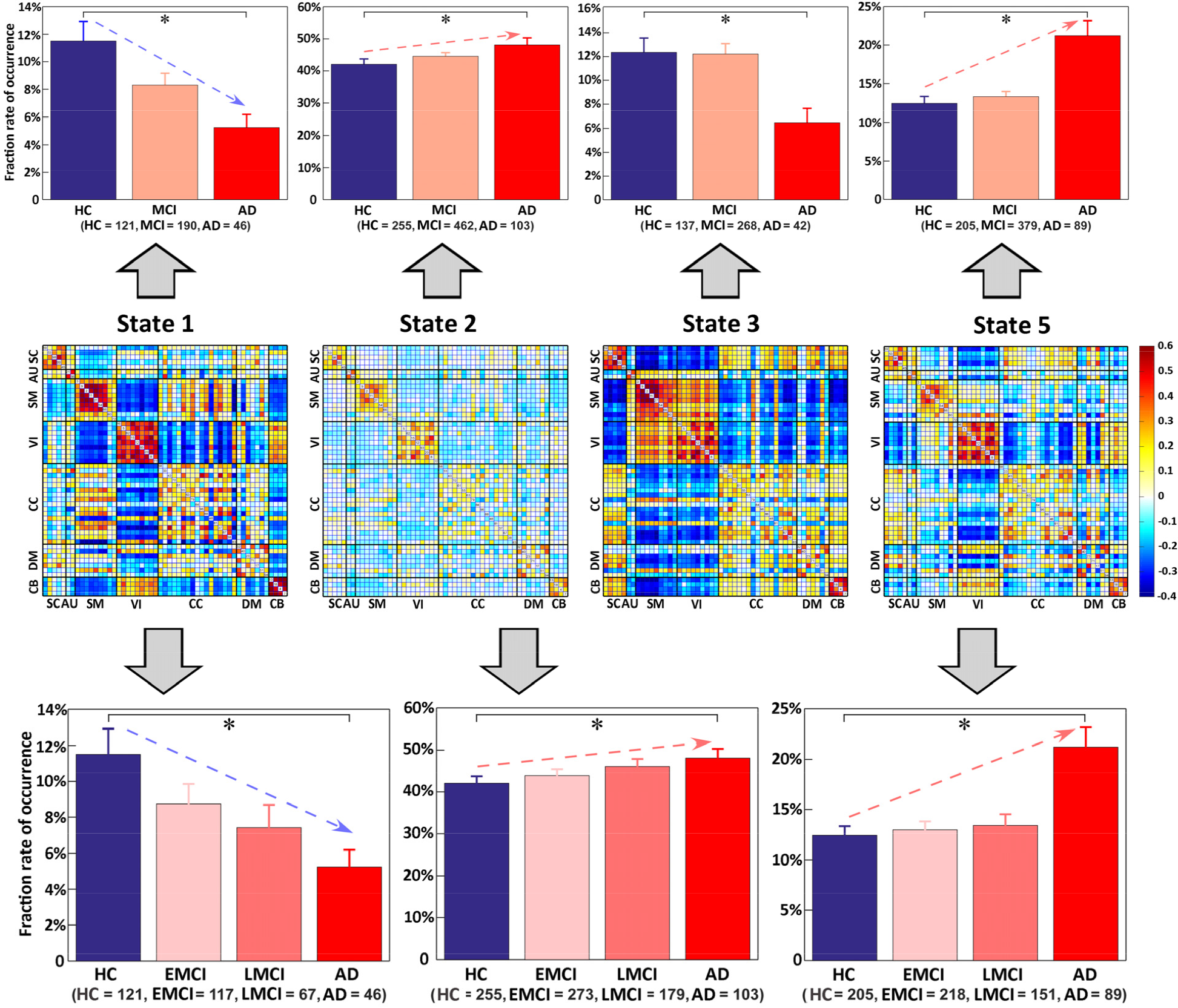
The results of study 3. The results revealed gradually changing patterns from healthy controls (HCs) to early MCI (EMCI) to late MCI (LMCI) to Alzheimer’s disease (AD), measured by dFNC measures. Upper: Group differences in the fraction rate of occurrences of dFNC states among HC, MCI, and AD. Middle: The discriminating dFNC states, along with the count of subjects that have at least one window clustered into the state. Bottom: Group differences in the fraction rate of occurrences of dFNC states among HC, EMCI, LMCI, and AD. Regarding the fraction rate of occurrences in each state, bar and error bar represent the mean and the standard error of mean, respectively. Significant group difference (false discovery rate corrected, *q* = 0.05) is indicated by asterisks.

### Study 4: Bipolar disorder (BD) and major depressive disorder (MDD) can be classified with high accuracy using brain spatial networks

In this study, we demonstrate that functional network measures derived using *NeuroMark* can be useful features for differentiating patients with challenging mental disorders. We focused on classifying bipolar disorder (BD) and major depressive disorder (MDD), both of which can exhibit strong depressive symptoms and are difficult to distinguish in clinical diagnosis. The features as input for the classification goal were ICNs computed using *NeuroMark* from fMRI data.

Fig. 6 shows the pipeline and evaluation result in differentiating the two groups (32 BD and 34 MDD patients). We applied an unbiased 10-fold cross-validation framework. Within the training datasets, the optimal ICN combination subset was selected from all functional networks of different functional domains and then was employed to train a support vector machine (SVM) classifier. After that, the testing datasets was classified using the built model and selected features. This resulted in a high mean of classification accuracy across 100 runs (overall accuracy: 91%, BD individual-class accuracy: 89%, MDD individual-class accuracy: 94%). Importantly, we observed that some networks (IC 56, IC 33, IC 40, IC 98, IC 80, and IC 20) frequently presented as features in all classification runs. These networks engaged middle temporal gyrus, insula, precuneus, putamen, superior parietal gyrus, and inferior occipital gyrus regions, suggesting they play a key role in separating BD and MDD.

**Fig. 6.**
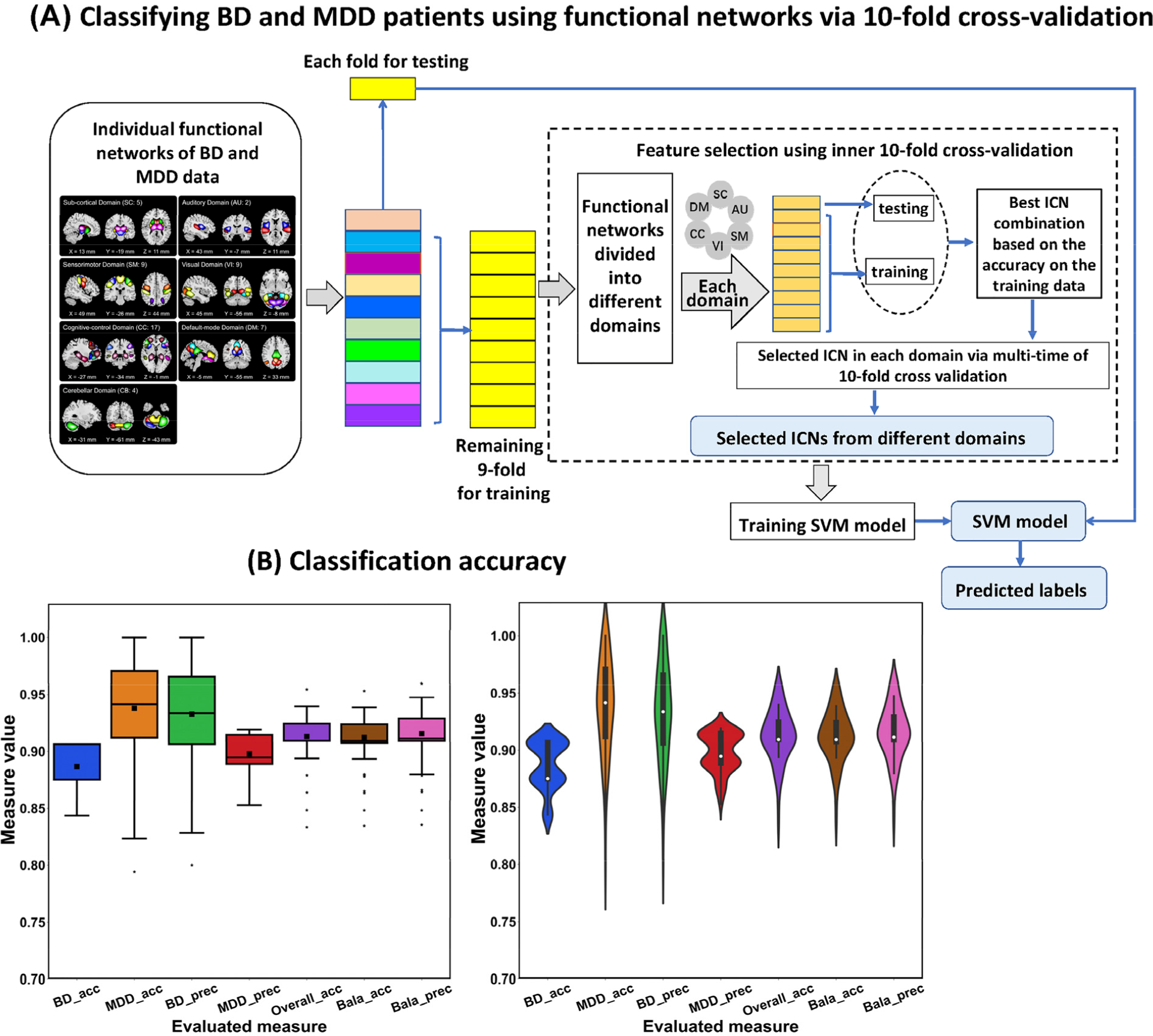
The classification pipeline and accuracy in study 4. (A) The pipeline of classifying between BD and MDD patients using spatial functional network maps as features, in which an unbiased 10-fold cross-validation procedure was applied. (B) The evaluated measures included individual-class accuracy (BD_acc and MDD_acc), individual-class precision (BD_Prec and MDD_prec), overall accuracy (Overall_acc), balanced accuracy (Bala_acc), and balanced precision (Bala_Prec). For each measure, we show the values from 100 runs using both boxplot and violinplot.

## Discussion

Clinical diagnosis of the neuropsychiatric disorders overwhelmingly relies on the pattern of symptoms. Neuroimaging measures may hold more objective, biology-based quantification of brain abnormalities and consequently provide additional biomarkers to guide diagnosis and treatment. The complexity of the brain requires the neuroscience community to analyze big-data samples assembled from multi-site studies to achieve statistically powerful findings. Exploring clinically related brain disorders via large-scale analysis to understand their underlying mechanisms and relationships also can help to redefine the disorder categories or develop new subtypes. Therefore, a standardized analysis method is greatly needed for working with big data in the neuroscience field.

Characterizing inter-relationship of spatially distributed brain regions using fMRI has been important for providing biomarkers in the neuroscience filed. Numerous methods for delineating and estimating functional network and connectivity have been proposed to promote the progress of precision medicine on brain disorders. ICA holds the promising ability to extract network features that retain more individual-level variability. However, due to the data-driven nature in ICA, synthesis across ICA-derived results presents a challenge that can hinder replication and cross-study comparison.

In this paper, we proposed a priori-driven ICA informed by reliable network templates to achieve linked analyses among different datasets, studies and disorders. In the method, spatial network priors can be leveraged by first estimating independent replicated network templates in order to automatically maintain subject correspondence, while also achieving individual variances by optimizing the individual-level network independence (Du and Fan, 2013; Lin et al., 2009). Here, the network templates were generated using two large-sample HC populations (including 1828 subjects) so as to minimize bias to any specific datasets to be analyzed. The use of such prior information can greatly reduce the search space and improve the likelihood of detecting useful markers (Cohen et al., 2017).

To assess the efficacy of *NeuroMark*, we performed four studies employing a large sample of fMRI data from 2454 subjects relating to six brain disorders to evaluate the method from different angles. The results clearly support that *NeuroMark* worked well in finding reproducible network markers of SZ from different datasets; disclosed the shared impairments in brain network connectivity between SZ and ASD by linking separate studies, revealed progressively changed subtle alterations from HC to MCI and to AD, and successfully distinguished brain disorders with similar symptoms (i.e. BP and MDD) based on brain functional network features.

In study 1, using our method, the SZ related changes showed a similar pattern in the functional network connectivity between two fully independent datasets. The primary overlapping networks included decreased connectivity between sub-cortical regions and cerebellum as well as increased connectivity between sub-cortical and auditory (or sensorimotor) regions. Our finding indicates that sub-cortical regions playing an important role in cognitive, affective, and social functions (Koshiyama et al., 2018), primarily the thalamus and caudate involved, are greatly affected in SZ. The thalamus is known for its important roles in visual, auditory, motor activity, emotion, memory and sensorimotor association functions, and the caudate nucleus integrates spatial information with motor behavior formulation. There has been evidence supporting functional abnormalities of these regions in SZ (Chen et al., 2019; Kirino et al., 2019; Woodward et al., 2012). Chen et al (Chen et al., 2019) revealed that thalamic hyperconnectivity with sensorimotor areas is related to the severity of cognitive deficits and clinical symptoms. In addition, they also showed the decreased interaction between thalamic and cerebellar regions. While our work found more alterations in SZ relative to the previous work, such as the enhanced connection between auditory and sub-cortical regions, we also confirmed that these impairments are replicated across different datasets. The overall results highlight the potential of *NeuroMark* as a tool for capturing reproducible biomarkers.

Study 2 shows that *NeuroMark* provides a way to link independent studies for pushing forward the understanding of related brain disorders. By comparing group differences between HC and ASD patients with the results from study 1, we found that the sub-cortical regions were significantly involved in both SZ and ASD. Specifically, we observed, for both disorders, decreased FNC between the cerebellum and thalamus (or caudate) and increased connectivity between the superior temporal gyrus and sub-cortical regions and between postcentral gyrus and thalamus. One previous study (Cerliani et al., 2015) using ICA-based fMRI analysis showed increased connectivity between networks encompassing basal ganglia and thalamus with the sensorimotor and auditory networks in ASD. Another recent study (Maximo and Kana, 2019) also reported cortico-subcortical functional connectivity differences. While our findings are consistent with previous work, this paper directly shows, for the first time, the overlap between SZ and ASD with respect to these changes. It is worth noting that although SZ and ASD are currently conceptualized as distinct disorders, there is overlap in symptoms such as social withdrawal and communication impairment (Ford et al., 2017). Historically, schizophrenia and autism were even once considered to be the same disorder expressed at different developmental periods. Our study provides biological evidence hinting at possible underlying mechanisms.

Study 3 showed that our method can identify subtle difference between disorders which manifest symptom severity along a continuous spectrum (taking MCI and AD for instance here). In this study, we used an advanced dynamic connectivity method, within the automated *NeuroMark* framework, to characterize brain function. Compared to HCs, AD patients exhibited more occurrence in weakly connected states, consistent with the results found in other brain disorders, including bipolar disorder (Rashid et al., 2014), schizophrenia (Damaraju et al., 2014; Du et al., 2016b), and autism (Fu et al., 2018). Importantly, MCI showed similar changing trends as AD, with a lesser degree than AD. These patterns position MCI as an intermediate stage between HC and AD. More interestingly, when we divided the MCI group into EMCI and LMCI, gradual changes were found in the dynamic features from HC to EMCI to LMCI to AD. The study showed that *NeuroMark* can capture subtle dFNC differences that help to characterize progression in cognitive impairment in dementia.

In study 4, we used the spatial functional network features extracted via the *NeuroMark* method to distinguish complex brain disorders with similar symptoms. Using unbiased 10-fold cross-validation, we achieved high classification accuracy (∼90%) between BD and MDD patients that had overlapping depressive symptoms and usually are difficult to be separated in clinical practice. The important discriminating brain regions involved the middle temporal gyrus, insula, precuneus, putamen, superior parietal and inferior occipital gyrus. Furthermore, the performance was better than a previous study (Osuch et al., 2018), even though in the previous work the group-level ICs were computed based on the MDD and BD data themselves.

In general, the *NeuroMark* method showed great promise. However, our approach has its limitations. One limitation of the method is that the present network templates were obtained only based on two independent datasets. The templates can be progressively improved and refined as more data are included, hopefully to generate functional network templates with greater reproducibility. In addition, the current network templates were estimated using a higher model-order (the number of ICs =100). In future, we will explore network estimation at different parcellation levels. Considering the ability to link different datasets, disorders and studies in our method, we also plan to provide a cloud computation platform that implements this approach. Our hope is that by using this robust method, functional network features can be widely studied and compared among numerous brain disorders.

A robust and fully automated ICA method was introduced in this paper for the analysis of fMRI datasets. The method can also be expanded to other modalities. Taking structural MRI for example, source-based morphometry (SBM) (Bergsland et al., 2018; Xu et al., 2009), a multivariate version of voxel-based morphometry (VBM), applies ICA to gray matter maps to detect common covariation among subjects and subject-associated weights. It is apparent that results of SBM vary across different datasets and runs. Using our method taking robust priors as guidance, the covariation patterns can be linked, thus resulting in comparable weights as features across different data (see regression based example here (Silva et al., 2014)). Group ICA is also useful for analyzing electroencephalography (EEG) data. Previous studies (Huster et al., 2015; Huster and Raud, 2018) extracted EEG sources by concatenating the data across the spatial dimension (see also the EEGIFT software: http://trendscenter.org/software). Generating robust a priori sources to guide the individual source computation will be an ongoing effort.

In summary, we present a method to generalize and standardize the calculation of possible brain imaging biomarkers that leverages the benefits of a data-driven approach to adapt to the individual data, while also providing comparability across multiple analyses. Results highlight the promise of the approach that we hope will be a useful stepping stone towards eventual application of such approaches in the clinic.

## Data Availability

2454 subjects were used, spanning six brain disorders (schizophrenia, autism spectrum disorder, depression, bipolar disorder, mild cognitive impairment and Alzheimer’s disease）. We have the agreements for the use of all data.

## Acknowledgements

This work was supported by National Natural Science Foundation of China (Grant No. 61703253 to YHD, 61773380 to JS), National Institutes of Health grants 5P20RR021938/P20GM103472 & R01EB020407 and National Science Foundation grant 1539067 (to VDC).

Data collection and sharing for Study 3 was funded by the Alzheimer’s Disease Neuroimaging Initiative (ADNI) (National Institutes of Health Grant U01 AG024904) and DOD ADNI (Department of Defense award number W81XWH-12-2-0012). ADNI is funded by the National Institute on Aging, the National Institute of Biomedical Imaging and Bioengineering, and through generous contributions from the following: AbbVie, Alzheimer’s Association; Alzheimer’s Drug Discovery Foundation; Araclon Biotech; BioClinica, Inc.; Biogen; Bristol-Myers Squibb Company; CereSpir, Inc.; Cogstate; Eisai Inc.; Elan Pharmaceuticals, Inc.; Eli Lilly and Company; EuroImmun; F. Hoffmann-La Roche Ltd and its affiliated company Genentech, Inc.; Fujirebio; GE Healthcare; IXICO Ltd.; Janssen Alzheimer Immunotherapy Research & Development, LLC.; Johnson & Johnson Pharmaceutical Research & Development LLC.; Lumosity; Lundbeck; Merck & Co., Inc.; Meso Scale Diagnostics, LLC.; NeuroRx Research; Neurotrack Technologies; Novartis Pharmaceuticals Corporation; Pfizer Inc.; Piramal Imaging; Servier; Takeda Pharmaceutical Company; and Transition Therapeutics. The Canadian Institutes of Health Research is providing funds to support ADNI clinical sites in Canada. Private sector contributions are facilitated by the Foundation for the National Institutes of Health (www.fnih.org). The grantee organization is the Northern California Institute for Research and Education, and the study is coordinated by the Alzheimer’s Therapeutic Research Institute at the University of Southern California. ADNI data are disseminated by the Laboratory for Neuro Imaging at the University of Southern California.

## Author Contributions

Yuhui Du proposed the whole analysis method and conducted the analyses on studies 1 and 2. Yuhui Du drafted the manuscript and prepared the figures. Zening Fu worked on study 3 and revised the paper. Jing Sui, Shuang Gao, Yuhui Du, and Ying Xing worked on study 4. Yuhui Du, Zening Fu, Dongdong Lin, Mustafa Salman, Anees Abrol, and Md Abdur Rahaman worked on the data preprocessing, network template extraction, and network measure computation. L Elliot Hong and Peter Kochunov collected the MPRC data and revised the paper. Jiayu Chen revised the paper. Vince Calhoun supervised the work and edited the paper. All authors have given final approval of this version of the article.

## STAR* Methods

### Lead contact and materials availability

Further information and requests for resources should be directed to and will be fulfilled by the Lead Contact, Yuhui Du.

### Method details

In the section, we first describe the proposed *NeuroMark*, and then introduce the four example studies to assess the capacity of the method.

### Approach for selecting data and computing brain mask

A rigorous criterion was implemented for subject selection to ensure high-quality data. For fMRI, we selected data with the properties: 1) data with head motions less than 3° rotations and 3 mm transitions along the whole scanning period; 2) data with more than 120 time points in fMRI acquisition; 3) data providing a successful normalization in the full brain. In terms of the third point, whether fMRI data have good normalization to the template is important in group ICA. We evaluated the normalization quality of data by comparing the individual-subject mask and the group mask. This method was applied to each study’s fMRI data separately. First, using the volume in the first time point, we calculated the individual mask for each subject by setting voxels showing greater values than 90% of the whole brain mean to 1. Next, we yielded a group mask by setting voxels included in more than 90% of the individual masks to 1. Then, for each subject, we calculated the correlations between the group mask and the individual mask. The correlations were calculated using voxels within the top 10 slices of the mask, within the bottom 10 slices of the mask, and within the whole mask, resulting in three correlation values for each subject. If a subject had correlations larger than the specified thresholds, we included this subject for further fMRI analysis. Finally, the group mask of each study was computed again based on the selected subjects’ individual masks.

### Identifying reliable functional network templates

The spatial network priors (i.e., the ICN templates) were obtained based on two independent HC datasets from the human connectome project (HCP, http://www.humanconnectomeproject.org/data/) and genomics superstruct project (GSP, https://www.nitrc.org/projects/gspdata). We preprocessed the GSP dataset using statistical parametric mapping (SPM12, http://www.fil.ion.ucl.ac.uk/spm/). Rigid body motion correction was performed to correct subject head motion, followed by the slice-timing correction to account for timing difference in slice acquisition. The fMRI data were subsequently warped into the standard Montreal Neurological Institute (MNI) space using an echo planar imaging (EPI) template and were slightly resampled to 3 × 3 × 3 mm^3^ isotropic voxels. The resampled fMRI images were further smoothed using a Gaussian kernel with a full width at half maximum (FWHM) = 6 mm. For the HCP dataset, we downloaded the preprocessed data from online and resliced them to the same spatial resolution (3 × 3 × 3 mm^3^) with the preprocessed GSP data using SPM12. More details in terms of the preprocessing on HCP data can be found online (http://www.humanconnectomeproject.org/data/). After quality control, in total 1005 individuals from the GSP dataset and 823 individuals from the HCP dataset were chosen (Table S1).

**Table S1.**
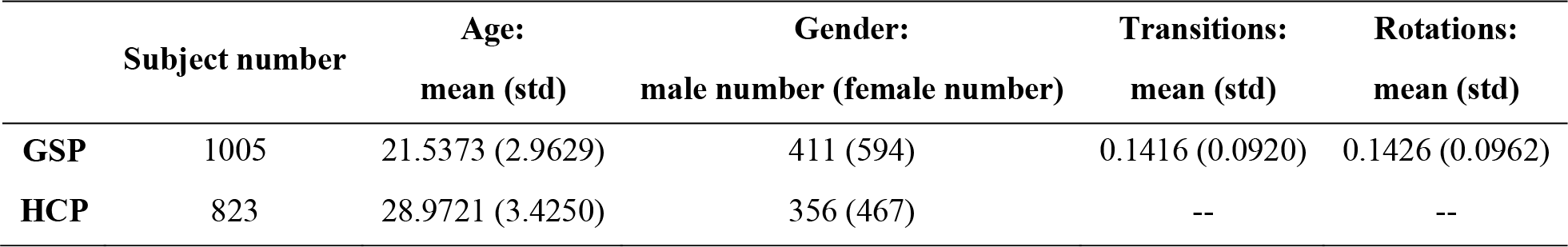
The demographic information of the GSP and HCP data

We conducted the following analysis for the GSP and HCP datasets, respectively, in order to obtain reliable network templates. Regarding each individual, principle component analysis (PCA) was first performed to reduce fMRI data to 110 principal components (PCs), which preserved more than 95% variance of the original data. Then, the individual-level PCs of each subject were concatenated across different subjects (1005 subjects for GSP or 823 subjects for HCP) and reduced into 100 PCs via another PCA at the group level. Next, the Infomax algorithm (Bell and Sejnowski, 1995) was used to decompose the 100 PCs into 100 ICs. This procedure was repeated 100 times using the ICASSO technique (Himberg and Hyvarinen, 2003), in which the best ICA run was selected to generate 100 reliable group-level ICs for each dataset (Ma et al., 2011).

We matched the two groups of ICs using a greedy spatial correlation analysis to find replicated networks. Here, a spatial similarity matrix ***C*** (size: 100 × 100) was obtained by computing the absolute value of Pearson correlation coefficients between spatial maps of ICs from GSP and that from HCP. Based on the matrix ***C***, the pair of ICs with the maximum correlation value were selected and considered as the first-matched components pair. If their original correlation value was negative, one of the ICs was sign-flipped. After identifying a matched ICs pair, the correlation values related to them in the matrix ***C*** were set to zero, resulting in a new similarity matrix ***C***_new_. As such, the matching procedure was repeated continually on the updated correlation matrices, until the final matched IC pair was found. IC pairs are considered to be reproducible if they show a higher spatial correlation than a given threshold 0.4, a more strict threshold than previous work (Smith et al., 2009). Next, we characterized a subset of these reproducible ICs as ICNs if they exhibited activation peaks in gray matter, had low spatial overlap with known vascular, ventricular, motion and other artifacts, and exhibited dominant low-frequency fluctuations in their TCs. Five fMRI experts carefully inspected those matched ICs, labeled meaningful ICNs and assigned them to different functional domains. ICs with more than three votes were identified as highly replicated ICNs. This resulted in two groups of highly similar ICNs from HCP and GSP dataset, respectively. Next, we selected the ICNs captured from the GSP dataset as the spatial network templates as they exhibited lower noise than the ICNs from the other group. Hereinafter, we use *N* to denote the number of network templates. The high reproduction degrees between networks of the two datasets are shown in Fig. S1.

**Fig. S1.**
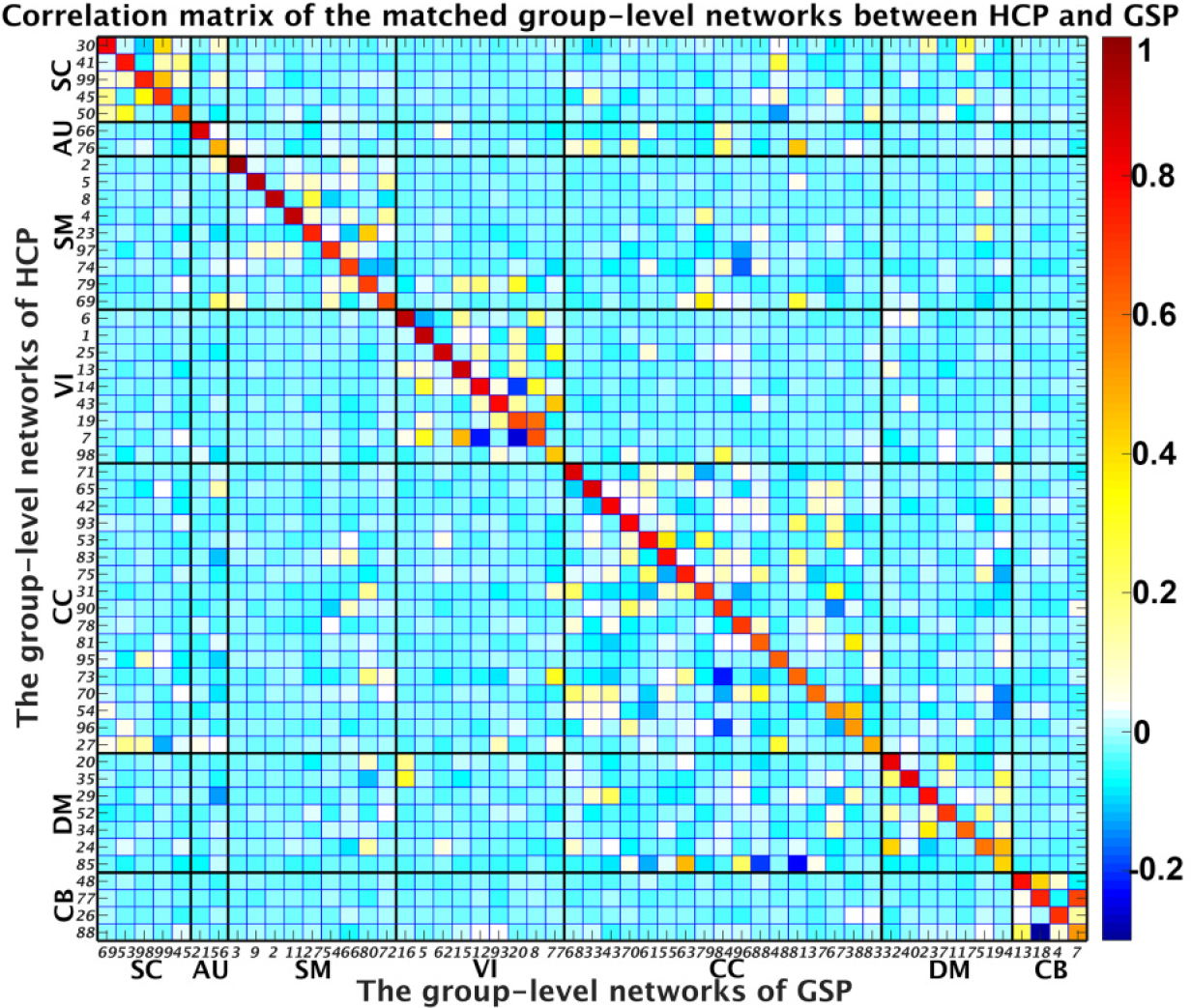
The sorted spatial correlation matrix between the matched two groups of functional networks. It is seen that the diagonal values are high, indicating the selected network templates are common between the GSP and HCP data.

### Estimating subject-specific functional brain networks

For each individual-subject fMRI data, ICNs are computed by Adaptive-ICA, an approach that automatically and adaptively estimates individual-level components using a prior define component templates. Two ICA algorithms (Du and Fan, 2013; Lin et al., 2010) available in the group ICA toolbox (GIFT) (http://trendscenter.org/software/) can be used for Adaptive-ICA. In this paper, we applied the GIG-ICA (Du and Fan, 2013) which uses a multiple-objective function optimization algorithm, by taking the obtained network templates and subject-specific fMRI data as input. Basically, there are two objective functions, one of which is to optimize the independence of networks in each subject’s fMRI data, while the other is to optimize the comparability between one subject-specific network and its related network template. In the original GIG-ICA algorithm (Du et al., 2016a; Du and Fan, 2013), group-level components used as guidance are computed from its own group data. Here, we use the labeled and ordered network templates validated from two independent datasets as spatial priors to estimate individual networks. The multiple-objective function represented in (1) was employed to compute one subject-specific network using a network template as guidance.

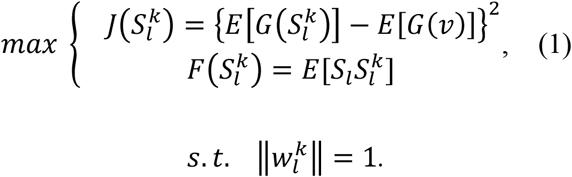

In (1), *S*_*l*_ denotes the *lth* network template, and 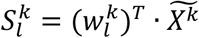 represents the estimated corresponding network of the *kth* subject, where 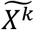 is the whitened *X*^*k*^ representing fMRI data matrix of the *kth* subject. Here, 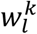 is the unmixing column vector, which is to be solved in the optimization functions. The first function is for optimizing the independence measure of 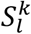, which is reflected using 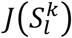, i.e., the negentropy of 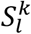. Here, *v* is a Gaussian variable with zero mean and unit variance; *G*(·) is a nonquadratic function. The second function 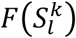 is used to measure the comparability between *S*_*l*_ and 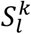. *E*[] denotes the expectation of variable. To solve the multiple-objective function optimization, a linear weighted sum method is applied to combine the two objective functions (Du and Fan, 2013). After the optimization, one subject-specific network can be obtained. Finally, for each subject, all *N* subject-specific networks corresponding to the *N* network templates and their relevant TCs are estimated from the data. Therefore, using this framework, all individual networks and their related TCs will not only be comparable across different datasets/studies/disorders as well as between previously analyzed data and new coming impendent data, but also show subject-unique characteristics in network activation and its temporal fluctuation.

### Functional brain network features

Using the proposed framework, multiple network features (in addition to the original ICNs and TCs) can be computed, including the interactions between functional networks, graph measures of functional organization, frequency information of networks’ fluctuations, as well as dynamic measures on both ICNs and their interactions. Taking network interaction as an example, static FNC (sFNC) can be obtained by computing the Pearson correlations between post-processed TCs of ICNs to yield a sFNC matrix reflecting the interaction between any two networks. Thus, each element of sFNC matrix represents the connectivity between a pair of functional networks. As such, while each ICN reflects intra-connectivity within brain functional network, sFNC matrix represents inter-connectivity strengths between different ICNs. Additionally, dynamic FNC (dFNC) can be investigated through a sliding time window approach (Allen et al., 2014; Hutchison et al., 2013), in which a tapered window obtained by convolving a rectangle with a Gaussian is often used to segment the entire TC of each ICN into several short TCs. For each window, the covariance matrix is then computed using the windowed TCs from different networks to measure the functional connectivity between ICNs within the window. To assess more accurate covariance matrix, a graphical LASSO method is usually applied to estimate the regularized inverse covariance matrix and then the covariance matrix from the inverse covariance matrix (Du et al., 2018). For each subject, the covariance matrix of each window can be concatenated to form an array (size: *N* × *N* × *T*, here *N* is the number of ICNs and *T* is the number of windows), representing the dynamic strength values of FNC along different windows.

### Studies for evaluating *NeuroMark*

#### Study 1: Investigating static functional network connectivity (sFNC) changes in schizophrenia (SZ): a replication study

In the first study, we assessed the ability of *NeuroMark* to identify reproducible disease biomarkers across different datasets. Here, the proposed method was implemented on two independent datasets, in order to detect brain changes of SZ patients relative to healthy controls. One dataset was from the Function Biomedical Informatics Research Network (FBIRN) including 210 SZ patients and 195 HCs. The other dataset was collected at the University of Maryland, Maryland Psychiatric Research Center (MPRC), including 251 SZ patients and 327 HCs. Resting-state fMRI data of SZ patients and HCs was preprocessed using the same preprocessing pipeline as for the GSP dataset, and then the same criteria were used for subject selection. We retained 137 SZ patients and 144 HCs in the FBIRN dataset, and 150 SZ patients and 238 HCs in the MPRC dataset for the further analysis.

Using the *N* ICN templates as priors, we performed GIG-ICA on each individual-subject fMRI dataset to extract spatial subject-level ICNs and their related TCs. Then, a sFNC matrix was obtained for each subject via computing Pearson correlation coefficients between the post-processed TCs. Specifically, in this paper we used the following steps to remove noise sources of TCs before sFNC computation, including 1) detrending linear, quadratic, and cubic trends; 2) conducting multiple regressions of the six realignment parameters and their temporal derivatives; 3) de-spiking to detect and remove outliers; and 4) band-pass filtering with [0.01-0.15] Hz. We averaged the individual sFNC matrices across subjects for the FBIRN and MPRC data, separately, to show if the connectivity patterns were comparable. Next, for each connection between two ICNs in the sFNC matrix, we investigated difference in connectivity strength between HCs and SZ patients by performing a two-tailed two-sample t-test (p < 0.05 with Bonferroni correction) for the FBIRN and MPRC datasets, separately, after regressing out age, gender and site effects. Finally, we investigated if the identified changes in SZ are reproducible between the two datasets by comparing their sFNC results from HC vs. SZ two-sample t-tests.

#### Study 2: Investigating the common static functional network connectivity (sFNC) alterations in autism spectrum disorder (ASD) and SZ: multi-study comparison

Since we computed the brain functional network templates using data from large-sample healthy population independent from the data being analyzed, it is feasible to link multiple independent studies. In study 2, we explored sFNC changes in ASD compared to HCs, and then evaluated the common impairments between SZ and ASD by summarizing the results from this analysis and study 1, each of that was performed independently of the other. We selected data from Autism Brain Imaging Data Exchange I (ABIDEI), provided by the National Institute of Mental Health. The ABIDEI dataset involves 17 sites and includes a total of 539 individuals with ASD and 573 age-matched HCs. We conducted preprocessing and subject selection using the same pipeline and criterion. After quality control, 398 ASD individuals and 471 HCs in the ABIDEI dataset remained.

Similar to study 2, using the *NeuroMark* method, we obtained *N* individual ICNs and then estimated their functional interaction (i.e. sFNC) for each subject. Furthermore, we investigated the HC vs. ASD differences on sFNC measures using two-tailed two-sample t-tests (p < 0.05 with Bonferroni correction). Since the features were extracted using a unified way by *NeuroMark*, we compared the symptom-related disorders (i.e. SZ and ASD) in terms of their overlapping alterations relative to HCs based on the results here and the results from study 1.

#### Study 3: Evaluating levels of cognitive performance/impairment linked to dynamic FNC

In this study, we investigated brain dynamic connectivity changes among Alzheimer’s disease (AD) patients, mild cognitive impairment (MCI) patients, and HCs. We used the publicly available Alzheimer’s Disease Neuroimaging Initiative (ADNI) dataset, which collected resting-state fMRI data from 275 HCs, 107 AD patients and 480 MCI patients. Using the same preprocessing and subject selection procedures, we had a total of 838 subjects (104 patients with AD, 470 patients with MCI, and 264 HCs) for analysis.

For each subject, we estimated dFNC using a sliding window approach (Allen et al., 2014). Since the datasets have different temporal resolutions, we performed interpolation on the TCs with longer TR to construct new TCs with the same nominal temporal resolution as those data with smallest TR and the same length of data. This procedure helps to control the potential impacts on the dynamic analysis caused by the different temporal resolutions. The tapered window was obtained by convolving a rectangle (window size = 40 TRs = 24.3 s) with a Gaussian (σ = 3) function. This window was slid in steps of 1 TR, resulting in total *T* = 468 windows for yielding dFNC matrices.

A K-means clustering analysis (Allen et al., 2014) was implemented on dFNC estimates to capture occurred states in time and across subjects. L1 norm was used as the distance function with the upper triangular (*N* × (*N* − 1)/2) values in the matrices as features. The optimal number of clusters was determined as five by the elbow criterion, which was within a reasonable range (4∼7) consistent with the previous dFNC studies on different brain disorders (Abrol et al., 2017; Du et al., 2018; Fu et al., 2019; Fu et al., 2018; Rashid et al., 2014).

Regarding each state, we computed its fraction rate of occurrence for each subject by computing the percentage of the number of time windows assigned to the state in the number of total windows. To investigate group differences in the fraction rate of each state, ANOVA was performed after regressing out age and gender. If the ANOVA resulted in a significant diagnosis effects, a generalized linear model (GLM) including age and gender was conducted to examine the group difference between any paired groups.

#### Study 4: Classification of two disorders with overlapping symptoms using spatial networks

In this study, we aimed to employ network features identified by the proposed approach for the classification of symptom-related disorders. Resting-state fMRI data from 32 patients with BD Type I and 34 patients with MDD were included. More details can be found in a previous study (Osuch et al., 2018). We conducted the preprocessing and subject selection using the same pipeline and criterion as other studies. Based on spatial network (i.e. ICN) features, we assessed the ability to distinguish the MDD and BD patients.

Using *NeuroMark*, the individual ICNs were estimated by taking the selected network templates as guidance after discarding cerebellum-related templates, due to cerebellum being partially missing in the scanned data. In order to evaluate if the ICNs can be powerful features to classify BD and MDD patients, we applied an unbiased 10-fold cross-validation framework, in which nine of ten folds were used as the training data and the remaining fold was used as the testing data successively. Consistent to the previous work (Osuch et al., 2018), we applied support vector machine with sigmoid kernel for classification. The feature selection and model training were performed only based on the training data.

Feature selection plays a key role in classification, especially for the high-dimensional network measures. In this work, we extracted the most discriminative ICN from each functional domain, and then combined the discriminative ICNs from all functional domains as features. In order to find the most discriminative ICN for each domain, we used an inner 10 times of 10-fold cross-validation procedure within the training set based on a forward ICN-selection technique. Basically, the ICNs were added one by one based on the classification accuracy on the inner testing data, evaluated using the model built using the inner training data. Then, for each run in the inner 10-fold procedure, the optimal ICN combination corresponding to the highest classification accuracy can be found. The ICN with the highest occurring frequency in the optimal ICN combination sets (across different repeats) was validated as the most-discriminative ICN for that domain. After that, the combined discriminative ICNs from different domains were used as features to train the outer training data. While the previous study (Osuch et al., 2018) that used group information from its own data, individual-level ICN features computed using our framework were more unbiased.

To quantify the classification results, we evaluated multiple measures including the individual class accuracy, individual class precision, overall accuracy, balanced accuracy and balanced precision (Cuadros-Rodriguez et al., 2016) based on the predicted and diagnosis labels. Different measures reflect the results from different angles. The individual class accuracy reported the ratio of correctly classified subjects of a particular class to the total number of subjects in the class. The individual class precision was defined as the number of correctly classified subjects of a particular class divided by the total number of subjects predicted as the class. The overall accuracy was computed as the ratio of correctly classified subjects of all classes to the total number of subjects of all classes. Additionally, we also computed the mean of individual class accuracies, called as the balanced accuracy. The individual class precision values were also averaged to represent the balanced precision. For each measure, we show the results from different repeats using both boxplot and violinplot.

